# Organised cancer screening among women who receive medically assisted reproduction treatments

**DOI:** 10.64898/2026.07.05.26357336

**Authors:** Adrian Raymond Walker, Signe Opdahl, Christos Venetis, Louisa Jorm, Neville F Hacker, Michael Chapman, Antoinette C Anazodo, Robert J Norman, Catharyn Stern, Ursula M Sansom-Daly, Georgina Mary Chambers, Claire Melissa Vajdic

## Abstract

There are no published data on cancer screening by women using medically assisted reproduction (MAR). Such data would aid interpretation of the cancer incidence and risk profiles for this group. Using linked population-based Australian health registries and administrative datasets, we compared organised publicly funded cervical and breast screening episodes for women who received one of three types of MAR and matched women who did not between 1991 and 2016. We modelled the proportion of women screened in the three years before and after first MAR treatment, adjusting for age, remoteness, parity, socio-economic disadvantage, cancer history, and uptake of the other screening program. After adjustment, a greater proportion of women who received MAR than women who did not had cervical screening before MAR (77.3%-84.1% vs 57.5%-62.0%, depending on treatment) and after MAR (77.0%-78.5% vs 68.1%-68.3%). Contrastingly, breast screening estimates were 7.6%-9.6% vs 9.3%-10.5% before MAR and 11.0%-15.0% vs 12.8%-14.9% after MAR.

---

The use of medically assisted reproduction (MAR) treatments, including assisted reproduction technology (ART) like in-vitro fertilisation (IVF) and intrauterine insemination (IUI) and ovulation stimulation (OS), is increasing worldwide.^1^ Due to the hormones used in many MAR treatments, there is concern these treatments may increase the development of cancer.^2–6^ Assessing this relationship using observational datasets is difficult, with likely multiple unmeasured confounding factors and the potential for detection bias.^4,7^ Given engagement with MAR services indicates a willingness to engage with healthcare services generally, women who receive MAR may be more likely to participate in organised cancer screening programs. If true, screening-based detection and treatment would prevent more preneoplastic, borderline and in-situ tumours from progressing to invasive cancers, but also increase the rate of detection of invasive cancers in MAR-treated women. This poses challenges for determining whether any change in the risk of cancer following MAR treatment is due to the treatment itself, or due to a difference in propensity for organised cancer screening. Although current Australian guidelines recommend women are up to date with cervical screening prior to attempting conception,^8^ there is no empirical evidence showing the rate of engagement with organised cancer screening by women who receive MAR.

The current study examined organised cervical and breast cancer screening episodes for women who received MAR and matched women who did not. We conducted two retrospective cohort studies of Australian women between 1994 and 2016.^9^ During this period, Australia offered a free or subsidised organised cervical screening program for women aged 18-70 years, and organised breast cancer screening programs were available free to women from age 40.^10,11^ Details of cohort formations are given in Supplementary Methods. Linked population-based registries and administrative health datasets were used to identify the cohorts and study variables.

We formed two MAR cohorts – a cervical cancer screening cohort of women aged 21 to 55 at first MAR treatment and up to four non-MAR controls (matched on birth year, parity at first MAR treatment date, age at first birth if one occurred, and remoteness across the available lifespan), in total 1,331,193 women, and a breast cancer screening cohort of women aged 43 to 55 at first MAR treatment and up to four matched non-MAR controls, in total 65,304 women. These age-ranges were selected to allow three years since women became age-eligible for each screening program. In each MAR cohort, we classified exposure based on the type of MAR treatment a woman first received: 1) ART, including IVF and ICSI; 2) Intrauterine insemination with follicle stimulating hormone or ART cycles cancelled before egg retrieval (IUI/OS); and 3) Dispensations of Clomiphene Citrate.

Tables S1 and S2 show the demographic profile of each cohort. The cervical cancer screening cohorts had median ages 31-35 years, and the breast cancer screening cohorts all had a median age of 44 years. For both cohorts, MAR-treated women were less socioeconomically disadvantaged than non-treated women.

We produced descriptive statistics on the proportion screened in each of the three years prior to first MAR treatment. We then modelled and predicted: 1) the proportion screened in the three years prior to first MAR treatment (or matched time for the non-treated women); 2) the number of days since the most recent screening in the three years prior to first MAR treatment; and 3) the proportion screened in the three years after first MAR treatment. We controlled for six variables during modelling: age at first MAR treatment; residential remoteness across the women’s lifespan as per the Accessibility/Remoteness Index of Australia Plus^12^; any children >20 weeks gestation prior to first MAR treatment; area-level Index of Relative Socioeconomic Disadvantage Percentile as per Socio-Economic Indexes for Areas (SEIFA)^13^; any history of notifiable cancer prior to first MAR treatment^14^; and participation in the alternative screening program (i.e., participated in organised cervical screening for the breast cancer cohort and vice versa) in the three years prior to first MAR treatment.

For all treatment types, a greater proportion of MAR-treated women underwent organised cervical cancer screening in each of the years preceding first MAR treatment compared to non-treated women (Figure 1). MAR-treated women also showed an increase in cervical screening the year immediately prior to MAR compared to the two years prior (from 34.4-37.5% to 45.7-49.8% depending on MAR treatment type) that was not seen in non-treated women. In contrast, compared to non-treated women, a consistently smaller proportion of women who received MAR attended organised breast cancer screening (noting very low screening rates of ∼3-4%).

**Figure 1:**
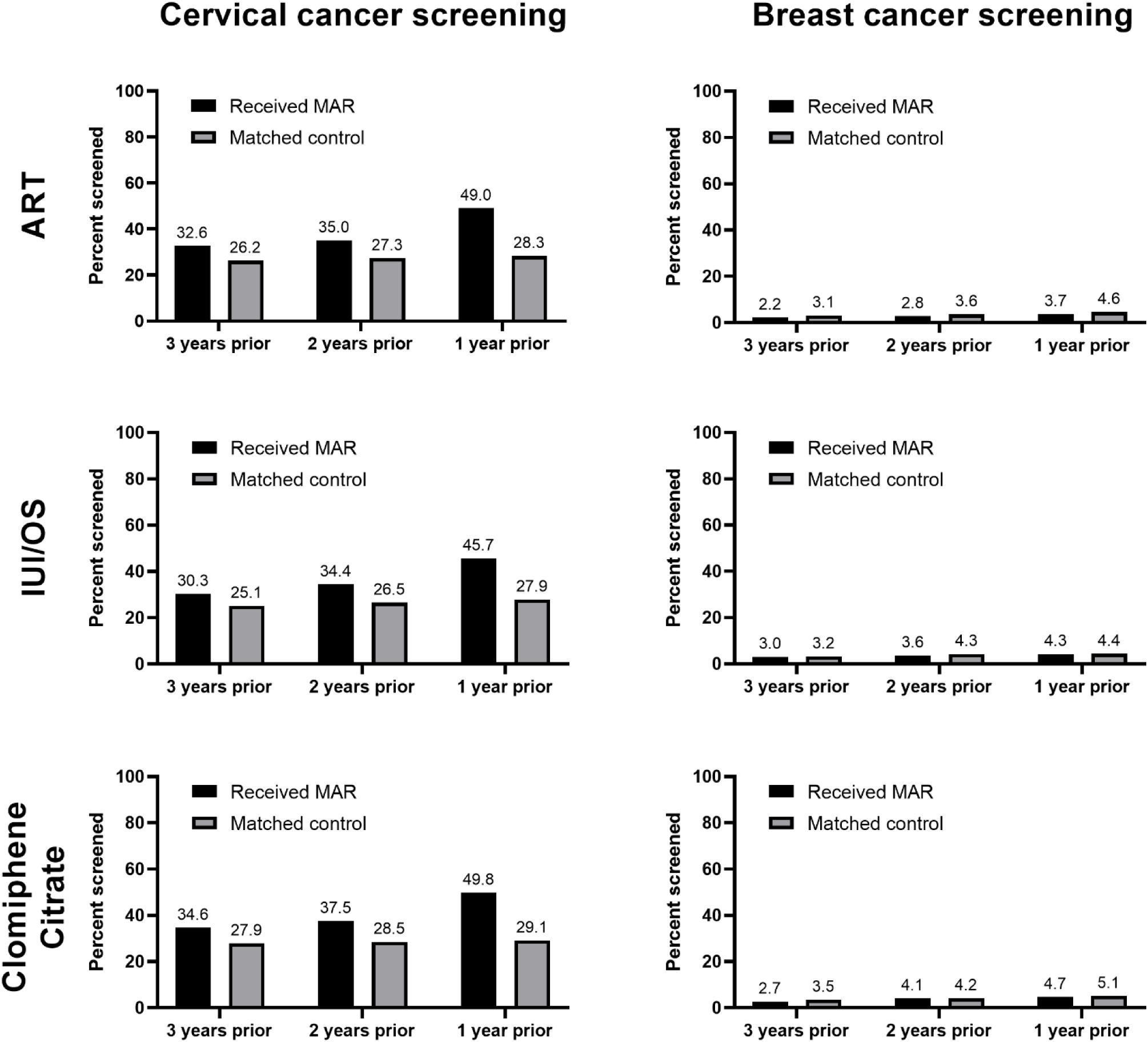
Proportion of the two screening cohorts screened for the relevant cancer each year prior to first medically assisted reproductive treatment (or matched date)

Table 1 shows the modelling predictions; results from all models can be seen at https://osf.io/nqkc2. For organised cervical cancer screening, 77.3-84.1% of women who received MAR were predicted to be screened in the three years prior to first MAR treatment, compared to 57.5-62.0% of non-treated women. Women who received MAR were predicted to have their most recent cervical cancer screen closer to the date of first MAR (334.9-339.2 days compared to 416.4-427.2 days). This pattern of cervical cancer screening was mirrored in the three years after MAR treatment, with 77.0-78.5% of women who received MAR being screened, compared to 68.1-69.5% of matched women.

**Table 1:** Model predictions for proportion screened in the three years prior/after receiving first medically assisted reproductive treatment (or matched date), and days since most recent screen before first medically assisted reproductive treatment.

| Cohort and screening type | Prediction after controlling for confounders | Assisted reproductive treatments |  | Intrauterine insemination/<br>Ovulation stimulation |  | Clomiphene Citrate |  |
| --- | --- | --- | --- | --- | --- | --- | --- |
|  |  | Estimate (95% CI) | Delta-method Standard Error | Estimate (95% CI) | Delta-method Standard Error | Estimate (95% CI) | Delta-method Standard Error |
| Cervical cancer |  |  |  |  |  |  |  |
|  | Proportion screened in three years prior to t0 |  |  |  |  |  |  |
|  | Women who received MAR | 81.6% (81.4%-81.8%) | 0.001 | 77.3% (76.9%-77.6%) | 0.002 | 84.1% (83.9%-84.3%) | 0.001 |
|  | Matched women | 59.2% (59.1%-59.4%) | 0.001 | 57.5% (57.2%-57.7%) | 0.001 | 62.0% (61.9%-62.2%) | 0.001 |
|  | Days since most recent screen in three years prior to t0 |  |  |  |  |  |  |
|  | Women who received MAR | 334.9 (333.4-336.5) | 0.805 | 340.9 (338.5-343.4) | 1.244 | 339.2 (337.7-340.7) | 0.779 |
|  | Matched women | 420.2 (419.0-421.3) | 0.575 | 416.4 (414.7-418.2) | 0.895 | 427.2 (426.2-428.3) | 0.536 |
|  | Proportion screened in three years after t0 |  |  |  |  |  |  |
|  | Women who received MAR | 77.2% (77.0%-77.5%) | 0.001 | 77.0% (76.6%-77.3%) | 0.002 | 78.5% (78.2%-78.7%) | 0.001 |
|  | Matched women | 68.3% (68.2%-68.5%) | 0.001 | 68.1% (67.9%-68.3%) | 0.001 | 69.5% (69.3%-69.6%) | 0.001 |
| Breast cancer |  |  |  |  |  |  |  |
|  | Proportion screened in three years prior to t0 |  |  |  |  |  |  |
|  | Women who received MAR | 7.6% (7.0%-8.2%) | 0.003 | 9.0% (8.0%-10.1%) | 0.005 | 9.6% (8.7%-10.5%) | 0.004 |
|  | Matched women | 9.3% (8.9%-9.6%) | 0.002 | 9.9% (9.3%-10.5%) | 0.003 | 10.5% (10.0%-10.9%) | 0.002 |
|  | Days since most recent screen in three years prior to t0 |  |  |  |  |  |  |
|  | Women who received MAR | 397.6 (373.2-422.0) | 12.472 | 410.8 (373.5-448.1) | 19.019 | 385.2 (357.6-412.8) | 14.082 |
|  | Matched women | 400.9 (388.1-413.7) | 6.539 | 416.1 (395.5-436.7) | 10.509 | 404.0 (388.4-419.6) | 7.971 |
|  | Proportion screened in three years after t0 |  |  |  |  |  |  |
|  | Women who received MAR | 11.0% (10.4%-11.7%) | 0.003 | 13.9% (12.8%-14.9%) | 0.006 | 15.0% (14.1%-15.9%) | 0.005 |
|  | Matched women | 12.8% (12.5%-13.2%) | 0.002 | 13.8% (13.2%-14.3%) | 0.003 | 14.9% (14.4%-15.4%) | 0.002 |

CI: Confidence interval; MAR: Medically Assisted Reproduction; t0: Time zero (first MAR treatment or matched date);

For organised breast cancer screening, 7.6-9.6% of women who received MAR were predicted to be screened in the three years prior to first MAR treatment, compared to 9.3-10.5% of non-treated women. Only women who received ART treatments had a significantly lower predicted proportion of breast cancer screening than non-treated women (7.6% [95%CI: 7.0%-8.2%] compared to 9.3% [95%CI: 8.9%-9.6%], Supplementary Model Output Table R1). The predicted days since most recent breast cancer screen prior to first MAR treatment were similar for the two groups (385.2-410.8 days in MAR-treated compared to 400.9-416.1 days in non-treated women). In the three years after first MAR treatment, 11.0-15.0% of women who received MAR treatment were predicted to receive breast cancer screening, compared to 12.8-14.9% of matched women. Women who received ART treatments had a lower predicted proportion of breast screening after MAR (11.0% [95%CI: 10.4%-11.7%] compared to 12.8% [95%CI: 12.5%-13.2%], Supplementary Model Output Table R3).

Our findings show organised cervical cancer screening is more common in women who received MAR compared to matched women who did not, both before and after treatment. Contrastingly, breast cancer screening was less common before and after ART but not for other MAR treatments. These screening patterns are broadly consistent with care aligned with national guidelines. Women are advised to be up-to-date with their cervical cancer screening prior to and after conception - in this case, conception via MAR treatment.^15,16^ On the other hand, organised mammographic breast cancer screening is contraindicated in women who are pregnant or seeking to conceive, and in women who are breastfeeding (e.g. BreastScreen NSW guidance).^17^

Population-level organised cervical screening programs reduce cervical cancer incidence.^18–21^ Given participation in cervical cancer screening is more common for women who receive MAR than their matched peers, it is not surprising that cervical cancer has a 40% lower incidence after MAR compared to the general population.^2^ The generally higher screening participation and increase just prior to MAR suggests screening is in response to requests by fertility specialists and general practitioners in women seeking to achieve pregnancy. Our evidence that 1-in-4 to 1-in-6 women were not up-to-date with cervical screening prior to MAR identifies missed opportunities for the prevention of cervical cancer during the clinical workup for MAR.

We had limited ability to examine uptake of organised breast screening by women who received MAR. During the study period, free organised breast screening was available for women from age 40, but women were only actively invited to attend the service from age 50. Women who take up free organised breast screening from age 40 are likely to be highly selected. In addition, because we required three years eligibility for breast screening prior to MAR, we were restricted to examining only women who first had MAR after age 43, a small subset (3.7%) of all MAR-treated women.

Our study benefited from access to near population-level capture of MAR treatment and organised breast and cervical cancer screening over a long time-period. However, we only had access to organised, publicly funded breast cancer screening episodes of care. It is estimated around 1-in-9 Australian women attend private breast screening services, more so younger women,^22^ but this service data are not collected at a population level. It is conceivable women having MAR are more likely to attend private services, due to their higher socioeconomic means. We were also only able to adjust for area-level and not person-level socio-economic status, a factor associated with both MAR and screening uptake.^23^

Overall, women who receive MAR may not have higher preventive healthcare engagement, but as expected, they do have greater participation in organised cervical screening, and this may be an indicator of greater engagement with reproductive health services, especially those that can be performed without referral to another service. For studies examining the relationship between MAR treatment and cancer, nuance is necessary when discussing how engagement with preventive healthcare services like organised cancer screening may be affecting findings.

## DATA AVAILABILITY

The data underlying this article cannot be shared for privacy reasons. The analyses were based on data from government registries and administrative (claims) datasets (Australian Department of Health and Aged Care, State and Territory Health Departments, and the Australian Institute of Health and Welfare). The data can be made available on request to each of the data custodians after ethical approval from the relevant Human Research Ethics Committees. The statistical analysis plan is available at https://osf.io/c6vmu, and analysis code at https://osf.io/nqkc2/.

## ACKNOWLEDGEMENTS

We acknowledge and thank the staff at the Australian Institute of Health and Welfare Data Linkage Unit, the Data Access and Assurance Unit, Data Services Branch, Queensland Health, the NSW Centre for Health Record Linkage, the Tasmanian Data Linkage Unit, the Centre for Victorian Data Linkage, and the Linkage, Data Engineering Outputs and ISPD Client Services teams at Western Australia Data Linkage Services for supporting the project and undertaking data linkage. We thank the Australian Institute of Health and Welfare and the population-based cancer registries of New South Wales (NSW), Victoria, Queensland, Western Australia, South Australia, Tasmania, the Australian Capital Territory (ACT) and the Northern Territory for the provision of data from the Australian Cancer Database. We also thank the following data custodians for providing the datasets used for this project: Australian Department of Health and Aged Care (Medicare Benefits Schedule, Pharmaceutical Benefits Scheme); Australian Institute of Health and Welfare (National Death Index); the NSW Ministry of Health, the Department of Health Victoria, the Data Services Branch Queensland Health, the Department of Health Western Australia, Preventive Health South Australia, the Department of Health Tasmania, HealthInfo ACT (perinatal data including the Midwives Notification System dataset); the jurisdictional Registries of Births, Deaths and Marriages (birth data); and BreastScreen Victoria, BreastScreen NSW, BreastScreen Queensland, BreastScreen WA, BreastScreen Tasmania and BreastScreen SA. We are grateful to the Victorian Consultative Council on Obstetric and Paediatric Mortality and Morbidity (CCOPMM) for providing access to the data used for this project and for the assistance of the staff at Safer Care Victoria. The conclusions, findings, opinions and views or recommendations expressed in this paper are strictly those of the authors. They do not necessarily reflect those of CCOPMM.

