## Supplementary for "Organised cancer screening among women who receive medically assisted reproduction treatments"

### SUPPLEMENTARY METHODS

#### Design and setting

This was a retrospective cohort study set in Australia between 1994 and 2016.

#### Cohort definition and formation

##### *Data sources and linkage*

This analysis was based on a broader set of studies on MAR and cancer described in Vajdic et al.[1] and Walker et al.[2]. Briefly, the cohort was drawn from a linkage conducted by the Australian Institute of Health and Welfare (AIHW) together with State-based Data Linkage Units. The linkage spine was formed from the Medicare Enrolment File, a data collection of all people enrolled in Australia's universal public health insurance scheme.

All datasets probabilistically linked to the spine are described in Vajdic et al.,[1] eTable 1: the Medicare Enrolment File to identify eligible women and their residential location,[3] the Medicare Benefits Schedule data collection to identify relevant MAR services,[4] the Pharmaceutical Benefits Scheme data collection to identify dispensations of MAR-relevant drugs[5] the National Death Index to determine date of death,[6] the Australian Cancer Database to determine date and type of incident cancer,[7] and State Perinatal Data Collections and Registries of Births, Deaths, and Marriages from NSW, ACT, QLD, VIC, WA, and TAS to determine history of pregnancy.

This study additionally included the National Cancer Screening Register from which we identified date of organised cervical cancer screening (1 Jan 1991- 31 Dec 2019); and six State BreastScreen registries from which we identified date of organised breast cancer screening (NSW, 1 Jan 1988-31 Dec 2020; VIC, 1 Jan 1999-31 Dec 2022; QLD, 1 Jan 1991-11 Apr 2024; SA: 1 Jan 1989-31 Dec 2022; WA, 1 Jan 1991-31 Dec 2019; TAS, 2008-2020, noting precise TAS BreastScreen dates were not provided to researchers and were estimated from the data).

*Cohort construction*

Beginning with the cohort described in Walker et al.,[8] we first removed women who had a record of death before any recorded screening episode (Supplementary Figure 1). Then, all women residing in the ACT were excluded, as we did not have access to ACT BreastScreen data.

For this study, we only considered cancer screening in the immediate three years before and after first MAR treatment. This time interval was chosen because the recommended screening interval for both screening programs was two-yearly during the study period, and this would allow for (i) delays in screening availability and (ii) time from first consultation for MAR to receiving MAR (screening participation prior to MAR) and pregnancy after MAR (screening participation after MAR).

To allow for three years follow-up time after MAR, we excluded all women who were first treated after 2016. To allow for three years of lookback time, we restricted to women who received their first MAR treatment (and their matched controls) from 1994 onwards, as both cancer screening programs commenced in Australia in 1991. Further exclusions were made by state of residence to account for the earliest state-specific BreastScreen data made available for this project; from 1995 (Queensland), 2002 (Victoria), or 2008 (Tasmania).

Following these exclusions, we defined two cohorts: 1) A cervical cancer screening cohort; and 2) a breast cancer screening cohort. In the cervical cancer screening cohort, we excluded women aged under 21 at first MAR treatment, as the National Cervical Screening Program began at age 18 in the study period. We also excluded women with a record of invasive cervical cancer before first MAR treatment in the Australian Cancer Database. Likewise, for the breast cancer screening cohort, we excluded women aged under 43 at first MAR treatment, since the breast cancer screening program allowed women from the age of 40 to participate upon request, starting between 1991 and 1995, depending on the jurisdiction. We excluded women who had a history of breast cancer in the Australian Cancer Database prior to first MAR treatment. For breast cancer screening, we excluded those with a history of in-situ or invasive cancer, as in-situ breast cancers were recorded in the Australian Cancer Database from 2002.

**Variable definitions**

*Exposure definition*

Exposure to medically assisted reproduction was defined as in Vajdic et al.,[1] eTable 2. Briefly, three exposures were identified: 1) Assisted reproductive treatments (ART), including IVF and ICSI; 2) Intrauterine insemination with follicle stimulating hormone or ART cycles cancelled before egg retrieval (IUI/OS); 3) Dispensations of Clomiphene Citrate. Women were assigned to a treatment group if they had the relevant treatment codes in the Medicare Benefits Scheme or Pharmaceutical Benefits Scheme datasets in the first 28 days after study entry (as defined by AIHW during data linkage).

In the calculation of all time-related variables, the time of first MAR treatment for control women was the date of first MAR treatment for their matched exposed woman/women.

*Outcome definition*

Cervical cancer screening was defined as any episode of cervical screening with type "C" in the National Cervical Screening Program dataset, indicating a routine Pap smear (cytology).

Breast cancer screening was defined as any routine mammographic screening episode in a BreastScreen dataset.

The National Cervical Cancer Screening program changed from pap smears every two years to HPV testing every five years on the 1 December 2017. As we required at least three years of follow-up time to assess cervical screening after MAR, and we do not know the date of first exposure (only the year), in the analysis of cervical screening after first MAR treatment we excluded all women with a first exposure date after 2013. This step resulted in 279,445 women being dropped from the cervical cancer cohort (63,472 exposed) for this part of the analysis.

*Other variable definitions*

Confounding variables were defined as per Walker et al.[8] Three variables were defined based on the variables used to match women who received MAR to women who did not: age at first MAR treatment; residential remoteness across the women's lifespan (as per the Accessibility/Remoteness Index of Australia Plus);[9] and whether the woman had any children (> 20 weeks gestation) prior to first MAR treatment. The Medicare Enrolment File was used to define age at first MAR treatment, and remoteness across the woman's life span. The jurisdictional Perinatal Data Collections and Registries of Births, Deaths, and Marriages were used to define parity prior to first MAR treatment. We also defined the area-level indicator of Index of Relative Socioeconomic Disadvantage Percentile as per Socio-Economic Indexes for Areas (SEIFA)[10] for each woman based on residential address in the Medicare Enrolment File; and whether a woman had any history of notifiable cancer prior to first MAR treatment from the Australian Cancer Database.[7]

#### Statistical methods

*Cohort description*

We calculated descriptive statistics for all variables by exposure type and screening cohort. We report mean, standard deviation, median, and interquartile range for continuous variables, and frequency and proportion for binary/categorical variables.

*Cancer screening prior to MAR treatment*

All analyses for cervical and breast cancer screening were conducted separately on their respective cohorts. To first describe the pattern of screening uptake in each cohort, we first produced histograms plotting the proportion of individuals screened each year prior to first MAR treatment.

For each type of screening, we fit two logistic regressions predicting the occurrence of cancer screening at any point in the three years prior to first MAR treatment. The first regression included only variables used in cohort matching (age at first MAR treatment, remoteness across lifespan, and any children prior to first MAR treatment), and a variable indicating whether the individual received MAR. The second model also included IRSD percentile, history of notifiable cancer prior to first MAR treatment, and history of screening in the alternative program in the previous three years (i.e., history of breast cancer screening when cervical cancer screening was the outcome, and vice versa).

Following each model, we predicted conditional estimates (using the *margins* command in STATA) of the proportion of individuals expected to be screened, based on whether they received MAR treatment or not. These regressions and conditional proportional estimates were run for each type of MAR treatment.

We then estimated the average time since last screening episode within the three years to MAR treatment. To first control for disparity in the number of women who underwent screening, we fit a logistic regression to predict presence or absence of a cancer screening episode in the previous three years (controlling for exposure to MAR, age at first MAR treatment, remoteness across lifespan, and parity prior to first MAR treatment, IRSD percentile, history of cancer prior to first MAR treatment, and history of screening in the alternative program). We used the output of this regression to construct inverse probability weights to predict the likelihood of screening participation. We then fit a Poisson regression on those that had been screened, using the weights calculated in the previous step. Finally, we calculated conditional marginal estimates of time to screening (using the *margins* command in Stata), based on whether they received MAR treatment or not.

*Cancer screening after MAR treatment*

To assess the proportion of individuals screened in the three years after MAR, we again fit two logistic regressions predicting the occurrence of cancer screening in the three years prior to first MAR treatment. The variables included were the same as those described above for predicting cancer screening prior to MAR, with the addition in the second model of a variable indicating history of the same type of screening in the three years prior to MAR (e.g., if the outcome was cervical screening in the three years after MAR, a variable indicating cervical screening in the three years prior to MAR was included). In this analysis, we additionally excluded all women who died within the three years following first MAR treatment.

*Registration and analysis code*

This analysis protocol for this study is registered at https://osf.io/c6vmu/. All analysis code is available at https://osf.io/nqkc2/. SAS version 9.4 (SAS Institute), R version 4.4.2 (R Project for Statistical Computing), and Stata version 19 (StataCorp) were used to implement the analysis, conducted between April 2024 to November 2024.

**Ethical approval**

All relevant human research ethics committees (HRECs) including the AIHW HREC (EO2019/5/1061), approved this study. Researchers accessed only linked anonymised data under a waiver of informed consent.

**Reporting of results**

As model predictions from the less complex and more complex models were very similar (identical to 2 decimal places), we report only the results from the more complex model in this manuscript. Results from all models with annotation, including coefficients from prediction models, can be seen at https://osf.io/nqkc2.

#### SUPPLEMENTARY FIGURES

**Figure S1: Cohort construction flowchart. The initial cohort is drawn from Walker et al. [2]**

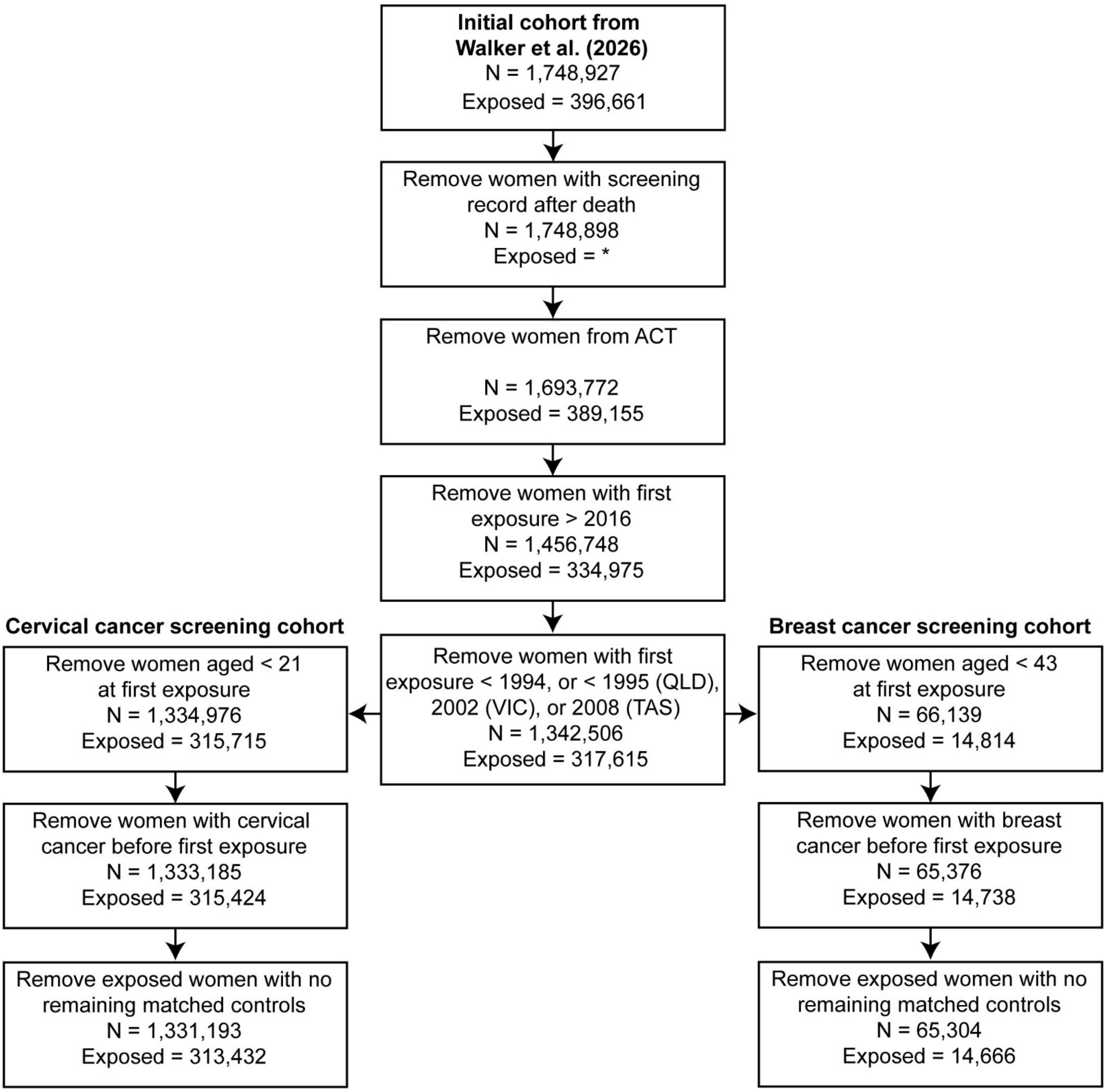

*Censored to prevent re-identification from small cell sizes
QLD = Queensland; VIC = Victoria; TAS = Tasmania

#### SUPPLEMENTARY TABLES

##### Table S1: Cohort demographics for cervical screening cohort.

| | Assisted reproductive treatments | | Intrauterine insemination/Ovulation stimulation | | Clomiphene citrate | |
|---|---|---|---|---|---|---|
| | **Treatment** | **Control** | **Treatment** | **Control** | **Treatment** | **Control** |
| **N** | 127,473 (23.7%) | 410,602 (76.3%) | 56,693 (24.6%) | 173,978 (75.4%) | 137,085 (23.0%) | 458,953 (77.0%) |
| **Age at study entry** | | | | | | |
| Mean (SD) | 34.8 (5.0) | 35.1 (5.0) | 34.2 (5.1) | 34.4 (5.0) | 31.9 (5.3) | 32.1 (5.3) |
| Median [IQR] | 35.0 [31.0 39.0] | 35.0 [32.0 39.0] | 34.0 [31.0 38.0] | 34.0 [31.0 38.0] | 31.0 [28.0 35.0] | 32.0 [28.0 36.0] |
| **Year of first MAR exposure** | | | | | | |
| 1994-1998 | 9,669 (7.6%) | 21,098 (5.1%) | 8,367 (14.8%) | 18,355 (10.6%) | NA | NA |
| 1999-2001 | 7,979 (6.3%) | 18,697 (4.6%) | 6,394 (11.3%) | 14,865 (8.5%) | NA | NA |
| 2002-2004 | 13,282 (10.4%) | 42,904 (10.4%) | 6,828 (12.0%) | 21,842 (12.6%) | 24,571 (17.9%) | 79,594 (17.3%) |
| 2005-2007 | 18,617 (14.6%) | 62,963 (15.3%) | 7,475 (13.2%) | 25,216 (14.5%) | 28,470 (20.8%) | 95,842 (20.9%) |
| 2008-2010 | 23,834 (18.7%) | 81,305 (19.8%) | 8,868 (15.6%) | 30,229 (17.4%) | 26,510 (19.3%) | 89,549 (19.5%) |
| 2011-2013 | 26,462 (20.8%) | 89,477 (21.8%) | 9,200 (16.2%) | 31,085 (17.9%) | 29,983 (21.9%) | 100,248 (21.8%) |
| 2014-2016 | 27,630 (21.7%) | 94,158 (22.9%) | 9,561 (16.9%) | 32,386 (18.6%) | 27,551 (20.1%) | 93,720 (20.4%) |
| **State** | | | | | | |
| New South Wales | 51,169 (40.1%) | 148,554 (36.2%) | 20,488 (36.1%) | 67,407 (38.7%) | 48,507 (35.4%) | 157,833 (34.4%) |
| Queensland | 20,761 (16.3%) | 88,058 (21.4%) | 16,564 (29.2%) | 36,782 (21.1%) | 30,304 (22.1%) | 93,587 (20.4%) |
| South Australia | 10,610 (8.3%) | 25,875 (6.3%) | 3,264 (5.8%) | 11,321 (6.5%) | 8,279 (6.0%) | 30,377 (6.6%) |
| Tasmania | 1,227 (1.0%) | 4,904 (1.2%) | 541 (1.0%) | 1,735 (1.0%) | 1,581 (1.2%) | 5,634 (1.2%) |
| Victoria | 31,219 (24.5%) | 100,732 (24.5%) | 8,639 (15.2%) | 38,552 (22.2%) | 40,001 (29.2%) | 123,662 (26.9%) |
| Western Australia | 12,487 (9.8%) | 42,479 (10.3%) | 7,197 (12.7%) | 18,181 (10.5%) | 8,413 (6.1%) | 47,860 (10.4%) |
| **Remoteness** | | | | | | |
| Major cities of Australia | 105,303 (82.6%) | 326,144 (79.4%) | 47,760 (84.2%) | 139,698 (80.3%) | 106,937 (78.0%) | 358,675 (78.2%) |
| Inner regional Australia | 14,368 (11.3%) | 57,332 (14.0%) | 6,242 (11.0%) | 24,049 (13.8%) | 19,955 (14.6%) | 65,751 (14.3%) |
| Outer regional Australia | 6,371 (5.0%) | 22,533 (5.5%) | 2,340 (4.1%) | 8,461 (4.9%) | 8,519 (6.2%) | 28,520 (6.2%) |
| Remote Australia | 987 (0.8%) | 2,960 (0.7%) | 246 (0.4%) | 1,186 (0.7%) | 1,208 (0.9%) | 3,926 (0.9%) |

| | | | | | | |
|---|---|---|---|---|---|---|
| Very remote Australia | 444 (0.3%) | 1,633 (0.4%) | 105 (0.2%) | 584 (0.3%) | 466 (0.3%) | 2,081 (0.5%) |
| **Index of Relative Socioeconomic Disadvantage quintile** | | | | | | |
| 1 | 15,247 (12.0%) | 60,751 (14.8%) | 6,405 (11.3%) | 25,258 (14.5%) | 23,236 (17.0%) | 71,139 (15.5%) |
| 2 | 16,989 (13.3%) | 62,990 (15.3%) | 7,068 (12.5%) | 26,509 (15.2%) | 21,566 (15.7%) | 74,221 (16.2%) |
| 3 | 23,960 (18.8%) | 79,494 (19.4%) | 11,161 (19.7%) | 33,820 (19.4%) | 28,507 (20.8%) | 91,339 (19.9%) |
| 4 | 27,555 (21.6%) | 86,738 (21.1%) | 13,177 (23.2%) | 36,623 (21.1%) | 28,521 (20.8%) | 95,832 (20.9%) |
| 5 | 43,722 (34.3%) | 120,629 (29.4%) | 18,882 (33.3%) | 51,768 (29.8%) | 35,255 (25.7%) | 126,422 (27.5%) |
| **Parity at first MAR exposure** | | | | | | |
| 0 | 101,311 (79.5%) | 284,865 (69.4%) | 44,781 (79.0%) | 123,106 (70.8%) | 94,121 (68.7%) | 286,026 (62.3%) |
| 1 | 16,082 (12.6%) | 51,754 (12.6%) | 8,612 (15.2%) | 22,989 (13.2%) | 30,627 (22.3%) | 83,238 (18.1%) |
| 2 | 5,853 (4.6%) | 42,729 (10.4%) | 2,214 (3.9%) | 16,341 (9.4%) | 8,197 (6.0%) | 55,277 (12.0%) |
| 3 | 2,668 (2.1%) | 19,176 (4.7%) | 691 (1.2%) | 7,203 (4.1%) | 2,555 (1.9%) | 22,343 (4.9%) |
| 4 | 1,038 (0.8%) | 7,356 (1.8%) | 246 (0.4%) | 2,654 (1.5%) | 906 (0.7%) | 7,828 (1.7%) |
| 5 and above | 521 (0.4%) | 4,722 (1.2%) | 149 (0.3%) | 1,685 (1.0%) | 679 (0.5%) | 4,241 (0.9%) |
| **Cancer prior to first MAR exposure** | | | | | | |
| Yes | 2,808 (2.2%) | 3,765 (0.9%) | 568 (1.0%) | 1,517 (0.9%) | 979 (0.7%) | 3,488 (0.8%) |
| **Cervical screen in 3 years prior to MAR** | | | | | | |
| Yes | 103,995 (81.6%) | 243,173 (59.2%) | 43,816 (77.3%) | 99,963 (57.5%) | 115,267 (84.1%) | 284,633 (62.0%) |
| **Cervical screen in 3 years after MAR** | | | | | | |
| Yes | 94,635 (74.2%) | 266,784 (65.0%) | 42,269 (74.6%) | 113,880 (65.5%) | 103,354 (75.4%) | 306,198 (66.7%) |
| **Breast screen in 3 years prior to MAR** | | | | | | |
| Yes | 1,025 (0.8%) | 4,017 (1.0%) | 466 (0.8%) | 1,471 (0.8%) | 665 (0.5%) | 2,556 (0.6%) |
| **Breast screen in 3 years after to MAR** | | | | | | |
| Yes | 2,827 (2.2%) | 10,112 (2.5%) | 1,289 (2.3%) | 3,703 (2.1%) | 1,961 (1.4%) | 6,366 (1.4%) |

MAR: Medically Assisted Reproduction

**Table S2: Cohort demographics for breast screening cohort.**

| | Assisted reproductive treatments | | Intrauterine insemination/Ovulation stimulation | | Clomiphene citrate | |
|---|---|---|---|---|---|---|
| | Treatment | Control | Treatment | Control | Treatment | Control |
| **N** | 7,605 (22.6%) | 26,091 (77.4%) | 2,976 (23.3%) | 9,800 (76.7%) | 4,500 (21.7%) | 16,200 (78.3%) |
| **Age at study entry** | | | | | | |
| Mean (SD) | 44.1 (1.4) | 44.1 (1.3) | 44.4 (1.6) | 44.4 (1.6) | 44.8 (2.2) | 44.8 (2.2) |
| Median [IQR] | 44.0 [43.0-45.0] | 44.0 [43.0-45.0] | 44.0 [43.0-45.0] | 44.0 [43.0-45.0] | 44.0 [43.0-46.0] | 44.0 [43.0-46.0] |
| **Year of first MAR exposure** | | | | | | |
| 1994-1998 | 442 (5.8%) | 1,003 (3.8%) | 334 (11.2%) | 741 (7.6%) | NA | NA |
| 1999-2001 | 354 (4.7%) | 888 (3.4%) | 299 (10.0%) | 732 (7.5%) | NA | NA |
| 2002-2004 | 725 (9.5%) | 2,405 (9.2%) | 354 (11.9%) | 1,176 (12.0%) | 817 (18.2%) | 2,777 (17.1%) |
| 2005-2007 | 1,130 (14.9%) | 3,975 (15.2%) | 367 (12.3%) | 1,300 (13.3%) | 1,051 (23.4%) | 3,724 (23.0%) |
| 2008-2010 | 1,512 (19.9%) | 5,410 (20.7%) | 418 (14.0%) | 1,506 (15.4%) | 895 (19.9%) | 3,290 (20.3%) |
| 2011-2013 | 1,596 (21.0%) | 5,725 (21.9%) | 580 (19.5%) | 2,082 (21.2%) | 896 (19.9%) | 3,298 (20.4%) |
| 2014-2016 | 1,846 (24.3%) | 6,685 (25.6%) | 624 (21.0%) | 2,263 (23.1%) | 841 (18.7%) | 3,111 (19.2%) |
| **State** | | | | | | |
| New South Wales | 3,390 (44.6%) | 9,414 (36.1%) | 1,180 (39.7%) | 3,654 (37.3%) | 1,777 (39.5%) | 5,670 (35.0%) |
| Queensland | 891 (11.7%) | 5,796 (22.2%) | 635 (21.3%) | 2,074 (21.2%) | 879 (19.5%) | 2,960 (18.3%) |
| South Australia | 551 (7.2%) | 1,611 (6.2%) | 173 (5.8%) | 699 (7.1%) | 204 (4.5%) | 1,282 (7.9%) |
| Tasmania | 60 (0.8%) | 385 (1.5%) | 32 (1.1%) | 107 (1.1%) | 41 (0.9%) | 183 (1.1%) |
| Victoria | 2,078 (27.3%) | 6,379 (24.4%) | 623 (20.9%) | 2,214 (22.6%) | 1,376 (30.6%) | 4,304 (26.6%) |
| Western Australia | 635 (8.3%) | 2,506 (9.6%) | 333 (11.2%) | 1,052 (10.7%) | 223 (5.0%) | 1,801 (11.1%) |
| **Remoteness** | | | | | | |
| Major cities of Australia | 6,670 (87.7%) | 20,021 (76.7%) | 2,559 (86.0%) | 7,637 (77.9%) | 3,667 (81.5%) | 12,424 (76.7%) |
| Inner regional Australia | 638 (8.4%) | 4,733 (18.1%) | 311 (10.5%) | 1,697 (17.3%) | 603 (13.4%) | 2,795 (17.3%) |
| Outer regional Australia | 259 (3.4%) | 1,139 (4.4%) | * | 379 (3.9%) | 198 (4.4%) | 824 (5.1%) |
| Remote Australia | 27 (0.4%) | 125 (0.5%) | * | 61 (0.6%) | 20 (0.4%) | 109 (0.7%) |
| Very remote Australia | 11 (0.1%) | 73 (0.3%) | * | 26 (0.3%) | 12 (0.3%) | 48 (0.3%) |
| **Index of Relative Socioeconomic Disadvantage quintile** | | | | | | |
| 1 | 742 (9.8%) | 4,160 (15.9%) | 340 (11.4%) | 1,509 (15.4%) | 896 (19.9%) | 2,476 (15.3%) |

| | | | | | | |
|---|---|---|---|---|---|---|
| 2 | 861 (11.3%) | 4,088 (15.7%) | 299 (10.0%) | 1,591 (16.2%) | 659 (14.6%) | 2,601 (16.1%) |
| 3 | 1,225 (16.1%) | 5,205 (19.9%) | 548 (18.4%) | 1,929 (19.7%) | 791 (17.6%) | 3,166 (19.5%) |
| 4 | 1,646 (21.6%) | 5,325 (20.4%) | 630 (21.2%) | 1,989 (20.3%) | 896 (19.9%) | 3,371 (20.8%) |
| 5 | 3,131 (41.2%) | 7,313 (28.0%) | 1,159 (38.9%) | 2,782 (28.4%) | 1,258 (28.0%) | 4,586 (28.3%) |
| **Parity at first MAR exposure** | | | | | | |
| 0 | 6,156 (80.9%) | 19,881 (76.2%) | 2,380 (80.0%) | 7,434 (75.9%) | 3,023 (67.2%) | 10,595 (65.4%) |
| 1 | 936 (12.3%) | 1,567 (6.0%) | 384 (12.9%) | 587 (6.0%) | 768 (17.1%) | 1,218 (7.5%) |
| 2 | 253 (3.3%) | 2,256 (8.6%) | 114 (3.8%) | 845 (8.6%) | 283 (6.3%) | 2,073 (12.8%) |
| 3 | 139 (1.8%) | 1,372 (5.3%) | 57 (1.9%) | 547 (5.6%) | 180 (4.0%) | 1,324 (8.2%) |
| 4 | 75 (1.0%) | 591 (2.3%) | 25 (0.8%) | 234 (2.4%) | 112 (2.5%) | 590 (3.6%) |
| 5 and above | 46 (0.6%) | 424 (1.6%) | 16 (0.5%) | 153 (1.6%) | 134 (3.0%) | 400 (2.5%) |
| **Cancer prior to first MAR exposure** | | | | | | |
| Yes | 97 (1.3%) | 402 (1.5%) | 33 (1.1%) | 159 (1.6%) | 64 (1.4%) | 281 (1.7%) |
| **Cervical screen in 3 years prior to MAR** | | | | | | |
| Yes | 6,229 (81.9%) | 14,990 (57.5%) | 2,346 (78.8%) | 5,550 (56.6%) | 3,740 (83.1%) | 9,941 (61.4%) |
| **Cervical screen in 3 years after MAR** | | | | | | |
| Yes | 5,361 (70.5%) | 14,658 (56.2%) | 2,156 (72.4%) | 5,585 (57.0%) | 3,229 (71.8%) | 9,467 (58.4%) |
| **Breast screen in 3 years prior to MAR** | | | | | | |
| Yes | 581 (7.6%) | 2,418 (9.3%) | 269 (9.0%) | 969 (9.9%) | 432 (9.6%) | 1,693 (10.5%) |
| **Breast screen in 3 years after to MAR** | | | | | | |
| Yes | 838 (11.0%) | 3,335 (12.8%) | 412 (13.8%) | 1,344 (13.7%) | 675 (15.0%) | 2,409 (14.9%) |

*Censored to prevent re-identification
MAR: Medically Assisted Reproduction

#### MODEL OUTPUT ANNOTATION FOR SCREENING DATA

##### Section purpose

This section outlines how to read the output from three excel spreadsheets ("Model output – ART exposure.xlsx", "Model output – IUI-OS exposure.xlsx", and "Model output – Clomiphene Citrate.xlsx") that report STATA model output to support the manuscript "Organised cancer screening in women who receive medically assisted reproduction treatment".

##### General structure

Each one of the three spreadsheets details output for one type of MAR exposure (as given in the name of the spreadsheet).

The three spreadsheets each have six tabs: two for each of the three research aims (R1, R2, R3, detailed below), with each research aim having separate output (tabs) for cervical cancer screening and breast cancer screening. The spreadsheets report direct model output from STATA.

The variables mentioned in the model output are detailed in the following table:

| Variable | Meaning |
|---|---|
| exposure | This variable codes for the exposure as a binary variable. "Case" is women who received MAR (compared to "Control", women who did not). |
| AgeT0 | This variable is the numeric variable of the women's age at first MAR treatment (or matched time for control women) |

| | |
|---|---|
| children_before_t0 | This variable codes if the women had a parity > 0 before first MAR treatment (or matched time for control women) |
| Remoteness | The ARIA+[9]index (as a category) of the woman at time of first MAR treatment (or matched time for control women). The category values are as follows:<br>Reference: Major Cities of Australia<br>1: Inner Regional Australia<br>2: Outer Regional Australia<br>3: Remote Australia<br>4: Very Remote Australia |
| cancer_before_t0 | This variable codes whether the woman had a notifiable cancer in the Australian Cancer Database |
| IRSD_Aus_percentile | This variable codes the SEIFA Index of Relative Socioeconomic Disadvantage[10] percentile for the women's location of residence at first MAR treatment (or matched time for control women) |
| cscreen_past_three/ bscreen_past_three | These two variables code whether the woman had an organised episode of cervical cancer screening (csreen) or breast cancer screening (bscreen) in the three years before first MAR treatment (or matched time for control women) |

**Research Question 1 (R1) – prediction of cohort screened before first MAR treatment**

These two tabs show the model output and marginal predictions on cancer screening in the three years prior to first MAR treatment (or matched time for control women). The "Basic" model includes only four variables (the exposure variable, and three confounders on which initial matching occurred for the cohorts, specifically age, parity and residential remoteness), and the "Complex" model includes all these variables plus the remainder listed in the table above. The raw logistic regression model output (from the *logit* command) is given on the left and the conditional marginal output (from the *margins* command) is given on the right. Margins are given separately for the women exposed to MAR ("Case") and for controls. These margins form the basis of some of the output presented in Table 1 of the manuscript.

**Research Question 2 (R2) – proportion of cohort screened before first MAR treatment and prediction of days to last screening**

These two tabs show both the proportion of the cohort screened prior to first MAR treatment, and the model output and marginal predictions for predicting time to most recent organised cancer screening.

The bar chart provided shows the raw proportion of the cohort who were screened in each of the years prior to first MAR treatment (or matched time for control women), split by whether the woman received MAR or not. This provides the basis for Figure 1 of the manuscript.

The "Selection" model output is generated from the logistic regression (using the *logit* command) for inverse probability of treatment weighting (predicting whether one received screening in the three years prior to first MAR treatment or matched time). The "Prediction" model output is generated from the weighted Poisson regression (using the *poisson* command) predicting the days to most recent cancer screening in the three years prior to first MAR treatment (or matched time for control women). On the right-hand side is the marginal predictions (generated using the *margins* command), first for the women exposed to MAR ("Case"), and then for controls. These margins form the basis of some of the output presented in Table 1 of the manuscript.

**Research Question 3 (R3) – prediction of cohort screened after first MAR treatment**

These two tabs show the model output and marginal predictions on cancer screening in the three years after first MAR treatment (or matched time for control women). The "Basic" model includes only four variables (the exposure variable, and three confounders on which initial matching occurred for the cohorts), and the "Complex" model includes all these variables plus the remainder listed in the table above. The raw logistic regression model output (from the *logit* command) is given on the left and the conditional marginal output (from the *margins* command) is given on the right. Margins are given separately for the women exposed to MAR ("Case") and for controls. These margins form the basis of some of the output presented in Table 1 of the manuscript.
